# The Effects of Education on Cognition in Older Age: Evidence from Genotyped Siblings^1^

**DOI:** 10.1101/2021.05.13.21257173

**Authors:** Jason Fletcher, Michael Topping, Fengyi Zheng, Qiongshi Lu

## Abstract

A growing literature has sought to tie educational attainment with later-life cognition and Alzheimer’s disease outcomes. This paper leverages sibling comparisons in educational attainment as well as genetic predictors (polygenic scores) for cognition, educational attainment, and Alzheimer’s disease to estimate effects of educational attainment on cognition in older age in the United Kingdom. We find that the effects of education on cognition are confounded by family background factors (∼40%) and by genetics (<10%). After adjustments, we continue to find large effects of education. College graduates have cognition scores that are approximately 0.75 SD higher than those who report no credentials. We also find evidence that educational effects on cognition are smaller for those with high polygenic scores for Alzheimer’s disease.

## Introduction

Cognitive health is one of the most critical aspects of health as people begin to enter the later stages of life (Evans et al., 2018). As individuals get older, they begin to face subtle changes to how they function and their overall cognitive ability. There is a great deal of variation in how cognition levels and trajectories vary in old age. Some people may experience and retain a high level of cognition in older age, whereas others in late life experience a decline (Gow et al., 2007). This has many implications because the cognitive ability that a person has in their older age has the potential to influence their autonomy, well-being, and quality of life (Díaz-Venegas et al., 2019). As populations continue to get older, it is imperative to look at specific indicators of cognition in later life, and consider how early life factors, such as educational attainment, continue to shape population-level cognition and health in old age.

There is a myriad of social, economic, and health-related consequences for older adults tied to their cognition. Failing cognition could lead to increased dependence on other people to complete tasks that were once simple (Gill et al., 2010), increased costs of potential treatments or adjustments to lifestyle practices (Vossius et al., 2011), and lead to disability, increased hospital visits, other morbidities and even death (Robertson, Savva, & Kenny, 2013). Research has shown that cognitive decline may be indicative for early phases of dementia or Alzheimer’s disease in later life, thus leading to greater consequences (Ritchie et al., 2016).

Cognition in later life is influenced by a multitude of predictors throughout many stages of the life course. Social connections that an individual has are an indicator of better cognitive function later in life, whereas social isolation is associated with many disadvantages (Evans et al., 2018; Kuiper et al., 2016). Level of cognition has also been positively influenced by lack of illness and mental health problems (Ritchie et al., 2016), greater amounts of exercise and physical activity throughout life (Sofi et al., 2011), being in a relationship with a significant other (Boss, Kang, & Branson, 2015), specific personality traits (Luchetti et al., 2016), and more favorable neighborhood contexts (Ailshire, Karraker, & Clarke, 2017). Conversely, cognition has been negatively associated with greater prevalence of illness (Ritchie et al., 2016), depression (van den Kommer et al., 2013; Zaninotto et al., 2018), lack of exercise or relationships (Sofi et al., 2011; Boss et al., 2015), disadvantaged neighborhood contexts (Ailshire et al., 2017), and age (Plassman et al. 2010). All these factors have implications not just for cognition, but other outcomes that involve health as well. However, despite all these factors that have the potential to influence cognition in later stages of the life course, one that gets the most attention is the role of education.

The influence of education in later life outcomes is well established in many disciplines and is the focus of many health outcomes (Galama, Lleras-Muney, & Kippersluis, 2018). People with higher levels of education tend to be far healthier on average, have greater longevity, and have fewer instances of morbidity throughout the life course (Davies et al., 2018; Lager & Torssander, 2012). This extends to the association that education has with cognition. Despite this, there is a debate about the relationship of education with many outcomes in later life, particularly if education influences these outcomes, or if the outcomes are a result of behavioral or genetic differences. While education has been shown to have positive effects on economic outcomes later in the life course, chiefly income (Clouston et al., 2012), there is a question of how strong the role is regarding cognition.

Research on educational attainment in early life and later cognition is vast (Chen et al. 2019; Clouston et al., 2012; Clouston et al., 2020; Lövdén et al., 2020; Schneewei, Skirbekk, & Winter-Ebmer, 2014; Wilson et al., 2019). Understanding this relationship is critical, since one’s cognitive ability plays a vital role in quality of life towards the end of the life course, and other outcomes such as lifestyle choices and social interactions (Langa et al., 2009; Ritchie et al., 2016). However, there have been some disputes regarding the determinants of cognition among adults in later life, particularly when it comes to the role of education. Some studies have shown that educational attainment is positively associated with cognition in later life (Richards & Hatch, 2011; Schneewei, et al., 2014; Yount, 2008). Chen and colleagues (2019) found that higher education among individuals in early life served as a defensive factor in aging, which in turn helped delay potential cognitive decline later on (Stern, 2009; Stern 2012). Conversely, other research has shown little to no link between education and cognitive abilities later in life (Glymour, Tzourio, & Dufouil, 2012; Zahodne, et al., 2011).

A major component of this debate in educational attainment’s role in cognition is due to whether education is a correlational or causal factor (Schneewei et al., 2014). For example, the association between these two factors could be a result of reverse causation, where high levels of cognitive ability in early life may influence educational attainment. However, education is something that could be influenced by a myriad of unobserved characteristics, which in turn could impact cognition in later life. A large literature has focused on the causal role education plays in childhood cognition, which ultimately could influence these same characteristics later in life (Clouston et al., 2020; Davies et al., 2018; Deary & Johnson, 2010; Richards & Sacker, 2011). Furthermore, there are several suggestions that higher levels of education in life not only affect cognitive ability but also may attenuate aging-associated declines in cognition, although there is doubt regarding the latter (Lövdén et al., 2020).

Research on policies and laws that are implemented at key points in the life course has been shown to have causal effects on later-life cognition. Banks and Mazzonna (2012) studied when compulsory school laws were implemented, specifically the age at which students could leave school. They found that educational attainment increases memory test scores in old age for students and, additionally, executive functioning in male students. Similarly, Glymour and colleagues (2008) discovered that increases in mandatory schooling of students led to improvements in performance on cognition tests, particularly memory scores, decades after they completed their schooling. However, they found that it may in fact not be causal due to other factors, such as genetics. Other studies have shown that higher educational attainment can aid in coping with age-related brain deterioration, which allows people to better handle cognitive tasks in later life (Lenehan et al., 2015).

In addition to issues of reverse causality, potential confounding between cognition and educational attainment has also been raised as an important empirical issue. One typical exploration of the role of confounding due to family background and genetics is to compare siblings. Data limitations have not allowed this technique in general, although some studies have examined the question in early life. These studies have shown that genetics appears to explain a large share of the similarity in cognition between siblings (Moorman, Carr, & Greenfield, 2018; Reynolds et al., 2014). Other research has studied sibling differences with regard to educational attainment and cognition but focused on how education is influenced by cognition, not the inverse (Polderman et al., 2015). Differential investments by parents may also lower levels of sibling similarity (Baier, 2019).^2^

This paper seeks to examine how sibling differences in educational attainment are related to differences in cognition in older age. This design, which is novel in the study of old age cognition, allows family background and shared genetics to be controlled. We add measured genetics that is unshared between siblings to the sibling difference design to further probe the possibility of genetic confounding between educational attainment and later-life cognition.

## Data

We used data from the UK Biobank (UKB) project. The participants, aged between 37 and 74 years, were originally recruited between 2006 and 2010.^3^ Cognition (fluid intelligence^4^) is measured in the UKB by summing the number of 13 logic puzzles that the participants could answer correctly in two minutes (Davies et al. 2018). This measure is only available for ∼200,000 of the 500,000 UKB participants, as it was added as a module toward the end of the recruitment window.

Educational levels of the UK Biobank participants were measured by mapping each major educational qualification that can be identified from the survey measures to an International Standard Classification of Education (ISCED) category and imputing a years-of-education equivalent for each ISCED category (Lee et al., 2018).

Although siblings are not identified in the survey, respondents’ genetics can be used to measure genetic relatedness among all pairs of respondents. We first use the UKB provided kinship file, listing all pairwise kinships among 100,000 pairs in the sample of nearly 500,000 individuals. We first choose pairs with kinship >0.2, which reflects first-degree biological relatives (parents/siblings). We then choose the remaining pairs who are <13 years apart in age, leaving ∼22,000 sibling dyads. We then chose to keep only one dyad from any family with more than one dyad, leaving ∼17,600 dyads. The number of dyads who also have non-missing cognition scores (only available for ∼1/3 of the full UKB sample) and educational attainment information is 4,138 (8,276 respondents). We include only respondents of European ancestry in our analysis.

We constructed polygenic scores (PGS) for three traits for which large genome-wide association studies (GWAS) are publicly available and do not contain UK Biobank samples: Alzheimer’s disease (Kunkle et al. 2019), cognition (Rietveld et al. 2014), and educational attainment (Lee et al., 2018). We clumped GWAS summary statistics by PLINK(42), using 1,000 Genome Project Phase III European samples as a reference for linkage disequilibrium (LD). We used an LD window size of 1Mb and a pairwise r^2^ threshold of 0.1. We did not apply any p-value thresholding to select variants (Choi & O’Reilly, 2019). PRSice-2 was used to calculate PGS. PGS of each trait was standardized to have a mean of zero and a standard deviation of one.

Table 1 presents descriptive statistics for our sample. Panel 1 shows the sample in the UK Biobank who have European ancestry, showing over 400,000 observations and 34,000 observations of siblings. Panel 2 limits this sample to those with a cognitive test available, where we have over 160,000 observations including 13,000 observations of individuals who have a sibling in the sample. Panel 3 restricts the sample to those sibling pairs where each member also has a cognitive test as well as educational attainment information available, providing nearly 8,300 observations. Comparing our analysis sample to the full sample, we see a high level of similarity among the variables in our analysis.

**Table 1.** Summary Statistics

| Variable | Obs | Mean | Std Dev | Min | Max |
| --- | --- | --- | --- | --- | --- |
| Sample: European Ancestry |  |  |  |  |  |
| Cognition | 164,150 | 6.3 | 2.1 | 0.0 | 13.0 |
| Education | 404,436 | 13.8 | 5.1 | 7.0 | 20.0 |
| EA-PGS (std) | 408,248 | 0.0 | 1.0 | -4.7 | 5.0 |
| AD-PGS (std) | 408,248 | 0.0 | 1.0 | -5.0 | 4.6 |
| Cog-PGS (std) | 408,248 | 0.0 | 1.0 | -4.9 | 4.8 |
| Age | 408,248 | 56.9 | 8.0 | 39.0 | 73.0 |
| Female | 408,248 | 0.5 | 0.5 | 0.0 | 1.0 |
| Family ID | 34,516 |  |  |  |  |
| Sample: Available Cognitive Test |  |  |  |  |  |
| Cognition | 162,812 | 6.3 | 2.1 | 0.0 | 13.0 |
| Education | 162,812 | 14.3 | 5.0 | 7.0 | 20.0 |
| EA-PGS (std) | 162,812 | 0.0 | 1.0 | -4.1 | 4.6 |
| AD-PGS (std) | 162,812 | 0.0 | 1.0 | -5.0 | 4.5 |
| Cog-PGS (std) | 162,812 | 0.0 | 1.0 | -4.9 | 4.3 |
| Age | 162,812 | 56.9 | 7.9 | 40.0 | 73.0 |
| Female | 162,812 | 0.5 | 0.5 | 0.0 | 1.0 |
| Family ID | 13,397 |  |  |  |  |
| Sample: Sibling Pairs |  |  |  |  |  |
| Cognition | 8,276 | 6.2 | 2.1 | 0.0 | 13.0 |
| Education | 8,276 | 13.8 | 5.0 | 7.0 | 20.0 |
| EA-PGS (std) | 8,276 | 0.0 | 1.0 | -3.7 | 3.9 |
| AD-PGS (std) | 8,276 | 0.0 | 1.0 | -3.7 | 4.0 |
| Cog-PGS (std) | 8,276 | 0.0 | 1.0 | -4.7 | 3.8 |
| Age | 8,276 | 57.5 | 7.2 | 40.0 | 70.0 |
| Female | 8,276 | 0.6 | 0.5 | 0.0 | 1.0 |
| Family ID | 8,276 |  |  |  |  |

Notes: Family ID refers to the family identification number of each respondent and is missing for those without biological siblings in the UKB.

## Results

Table 2 presents our main results. We include separate indicator variables for each educational category, with “no credential” as the omitted group. Column 1 shows the associations for the sample of respondents with European ancestry and non-missing cognition scores and allows a comparison with our analysis sample of siblings. The results are nearly identical and suggest large differences in cognition by educational category. For example, respondents with A-level credentials are over 2.3 points higher (1 SD) than those without credentials. Column 3 adds PGS for educational attainment, Alzheimer’s disease, and cognition. One standard deviation in the AD-PGS is associated with a 0.08 point reduction in cognition. Notably, these genetic controls only reduce the associations between educational attainment and cognition by ∼10%, suggesting a modest role of genetic confounding.

**Table 2.** Main Results

| Outcome<br>Sample<br>Fixed Effects? | Cognition<br>Full<br>None | Cognition<br>Pairs<br>None | Cognition<br>Pairs<br>None | Cognition<br>Pairs<br>Sibling | Cognition<br>Pairs<br>Sibling |
| --- | --- | --- | --- | --- | --- |
| Education = 10 | 1.392***<br>(0.016) | 1.394***<br>(0.065) | 1.352***<br>(0.065) | 0.753***<br>(0.091) | 0.737***<br>(0.091) |
| Education = 13 | 2.318***<br>(0.019) | 2.365***<br>(0.082) | 2.287***<br>(0.082) | 1.373***<br>(0.119) | 1.343***<br>(0.118) |
| Education = 15 | 1.450***<br>(0.024) | 1.450***<br>(0.103) | 1.407***<br>(0.103) | 0.948***<br>(0.133) | 0.940***<br>(0.133) |
| Education = 19 | 0.823***<br>(0.022) | 0.820***<br>(0.092) | 0.780***<br>(0.091) | 0.461***<br>(0.117) | 0.461***<br>(0.117) |
| Education = 20 | 2.743***<br>(0.015) | 2.710***<br>(0.065) | 2.600***<br>(0.066) | 1.535***<br>(0.104) | 1.493***<br>(0.104) |
| EA-PGS (std) |  |  | 0.138***<br>(0.023) |  | 0.136***<br>(0.042) |
| AD-PGS (std) |  |  | -0.083***<br>(0.021) |  | -0.082**<br>(0.037) |
| Cognition-PGS (std) |  |  | 0.065***<br>(0.023) |  | 0.055<br>(0.042) |
| Age | -0.003***<br>(0.001) | -0.000<br>(0.003) | -0.002<br>(0.003) | -0.013*<br>(0.007) | -0.012*<br>(0.007) |
| Female | -0.229***<br>(0.009) | -0.274***<br>(0.042) | -0.276***<br>(0.042) | -0.250***<br>(0.056) | -0.252***<br>(0.056) |
| Constant | 4.805***<br>(0.055) | 4.616***<br>(0.254) | 4.815***<br>(0.254) | 6.845***<br>(0.532) | 6.894***<br>(0.531) |
| Observations | 162,812 | 8,276 | 8,276 | 8,276 | 8,276 |
| R-squared | 0.207 | 0.212 | 0.220 | 0.069 | 0.074 |
| Number of Pairs |  |  |  | 4,138 | 4,138 |

Standard errors in parentheses. 20 PCs are controlled but not reported
\*\*\* p<0.01, \*\* p<0.05, \* p<0.1

Columns 4 and 5 present our sibling difference specifications. In general, the associations between educational attainment and cognition shrink by ∼40-50% compared to the results that do not control for sibling fixed effects; adding sibling differences in PGS to these models does not change these results. Overall, the results suggest a considerable level of confounding by family background and genetics but continue to point to remaining large associations between educational attainments and cognition in older age.

We next consider whether there is evidence for gene-environment interactions (GxE) between educational attainment (EA) and later-life cognition (Cog). Table 3 first explores the possibility of selection into education, which would suggest an important gene-environment correlation, where polygenic scores would predict both the “environment” of educational attainment and the old age cognition outcome of interest. Column 1 presents associations predicting educational attainment based on sibling differences in polygenic scores, with controls for age fixed effects, sex, and genetic principal components. Even between siblings, we do find evidence that EA-PGS predicts educational attainment, as found in other studies (Fletcher et al., 2020; Lee et al., 2018). In contrast, we find no association between Alzheimer’s disease (AD)-PGS and educational attainment, which mirrors earlier work focusing on *APOE* that suggests these variants do not impact decisions early in the life course (Cook & Fletcher, 2015). Thus, the AD-PGS appears to be a reasonable candidate to consider in GxE analysis.

**Table 3.**
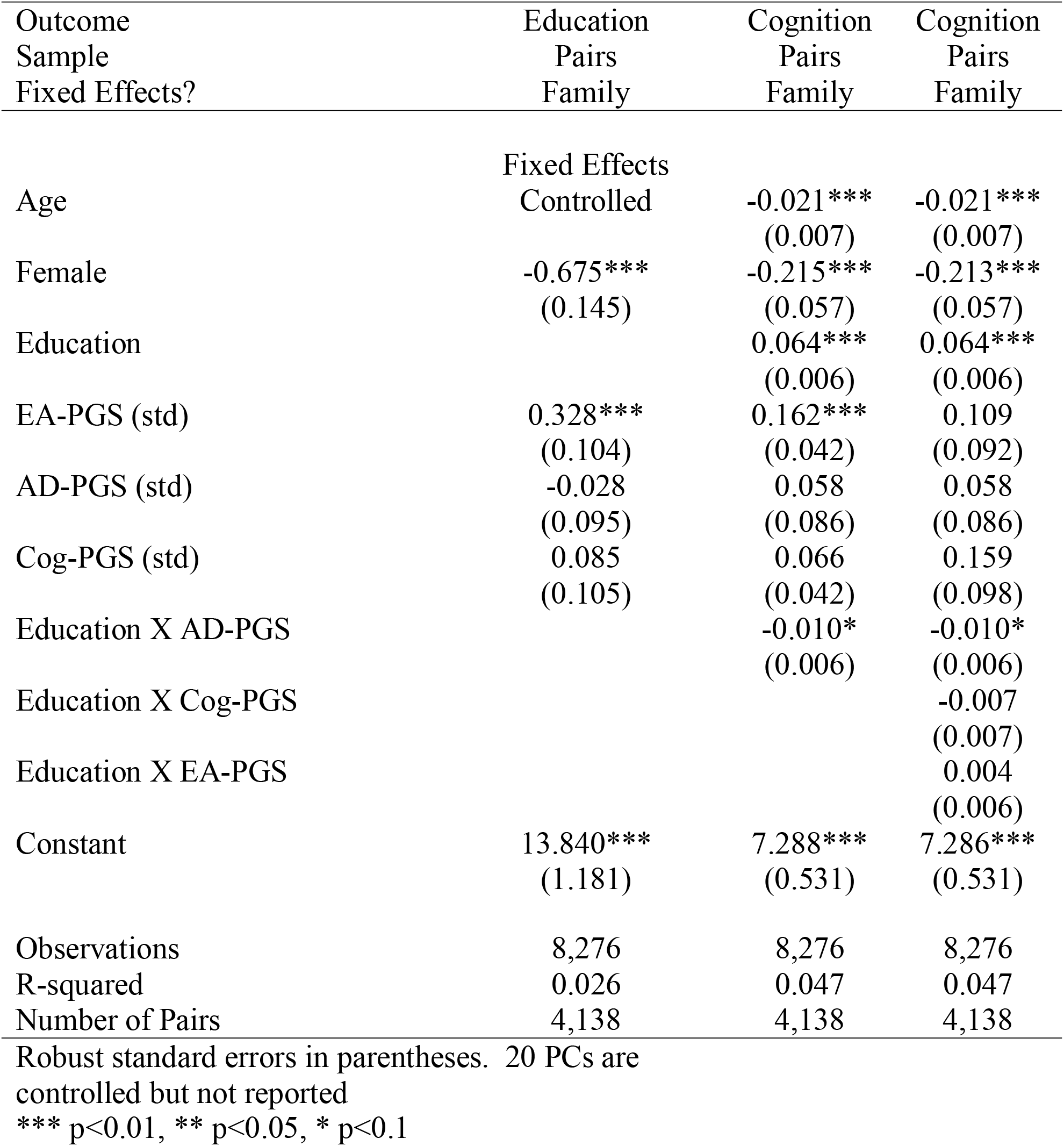
Selection and GxE

With the findings from Column 1 suggesting no evidence of gene-environment correlation for the AD-PGS, Columns 2 and 3 examines gene-environment interactions. The results show evidence that the impacts of educational attainment on later-life cognition are reduced for individuals with a higher genetic risk of Alzheimer’s disease (higher AD-PGS scores) and that this finding is not affected by also controlling for interactions between educational attainment and EA-PGS and Cog-PGS. In order to further explore these findings, Table 1A in the appendix examines gene-environment interactions for each level of educational attainment rather than treating attainment as a continuous measure. The results continue to show evidence that individuals with higher AD-PGS have smaller old age cognitive benefits from educational attainment than do individuals with lower AD-PGS.

## Discussion

Previous studies have sought to tie the role that educational attainment plays in cognition later in life. However, much of the literature has found mixed results, including positive and negative associations, with this relationship (Glymour et al., 2012; Richards & Hatch, 2011; Yount, 2008; Zahodne, et al., 2011). Additionally, research has focused attention on whether the relationship between education and cognition in later life is causal or correlational as a result of social, economic, behavioral, or genetic factors (Clouston et al., 2020; Davies et al., 2018; Schneewei et al., 2014). This paper applied a new research design to these questions by examining how sibling differences, as well as genetic predictors, influence the effects of educational attainment on cognition in old age. The analyses conducted indicate that there are indeed associations between educational attainment and cognition in older age, even controlling for confounding by family background and genetic factors.

There are important limitations to this study. First, the analysis is done with sibling dyads, which does not allow an analysis of individuals with no siblings. Another limitation is that this study does not include an examination of individuals of different racial and ethnic backgrounds. Future research should seek to examine how sibling and genetic differences impact the association between educational attainment and cognition by racial groups, as research has shown that there are racial differences at the population level in cognition and Alzheimer’s disease in later life (Barnes et al., 2015; Howell et al., 2017). A third limitation is the lack of experimental or quasi-experimental variation in educational attainments. We instead leverage sibling differences in educational attainment (controlling for genetic factors) to examine sibling differences in cognition. A fourth limitation is that only a subsample of the UKB has information on cognition, which reduced the power of our analysis, although we still use one of the largest datasets of siblings that measure genetics and cognition in older age.

Despite the limitations outlined above, this research is the first to use sibling fixed-effect models and genetic predictors to estimate the effects of education on cognition in older age. Results point to important associations between education and cognition in older age. These findings largely support previous work done on how education can aid in coping with age-related deterioration (Lenehan et al., 2015), and thus act as a vehicle to maintain or improve cognition in later life. We also provide novel evidence of an interaction between educational attainment and the PGS for AD in predicting cognition in later life, suggesting those at higher genetic risks receive smaller benefits from education than those at lower genetic risks. This highlights the need to examine the intersection of both social and genetic factors when studying outcomes in late life.

## Data Availability

Data available from UK Biobank

---

**Table 1A:**
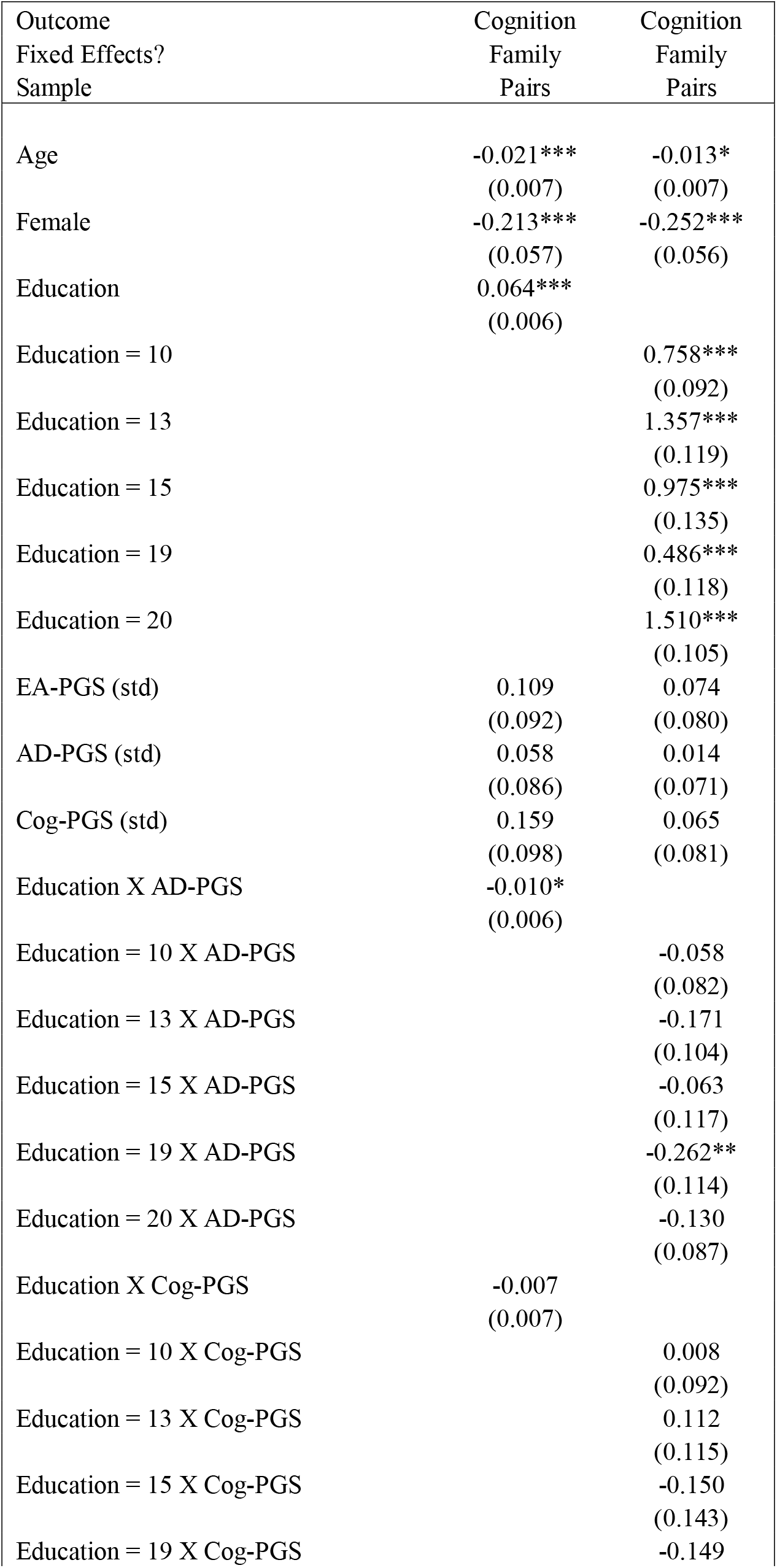
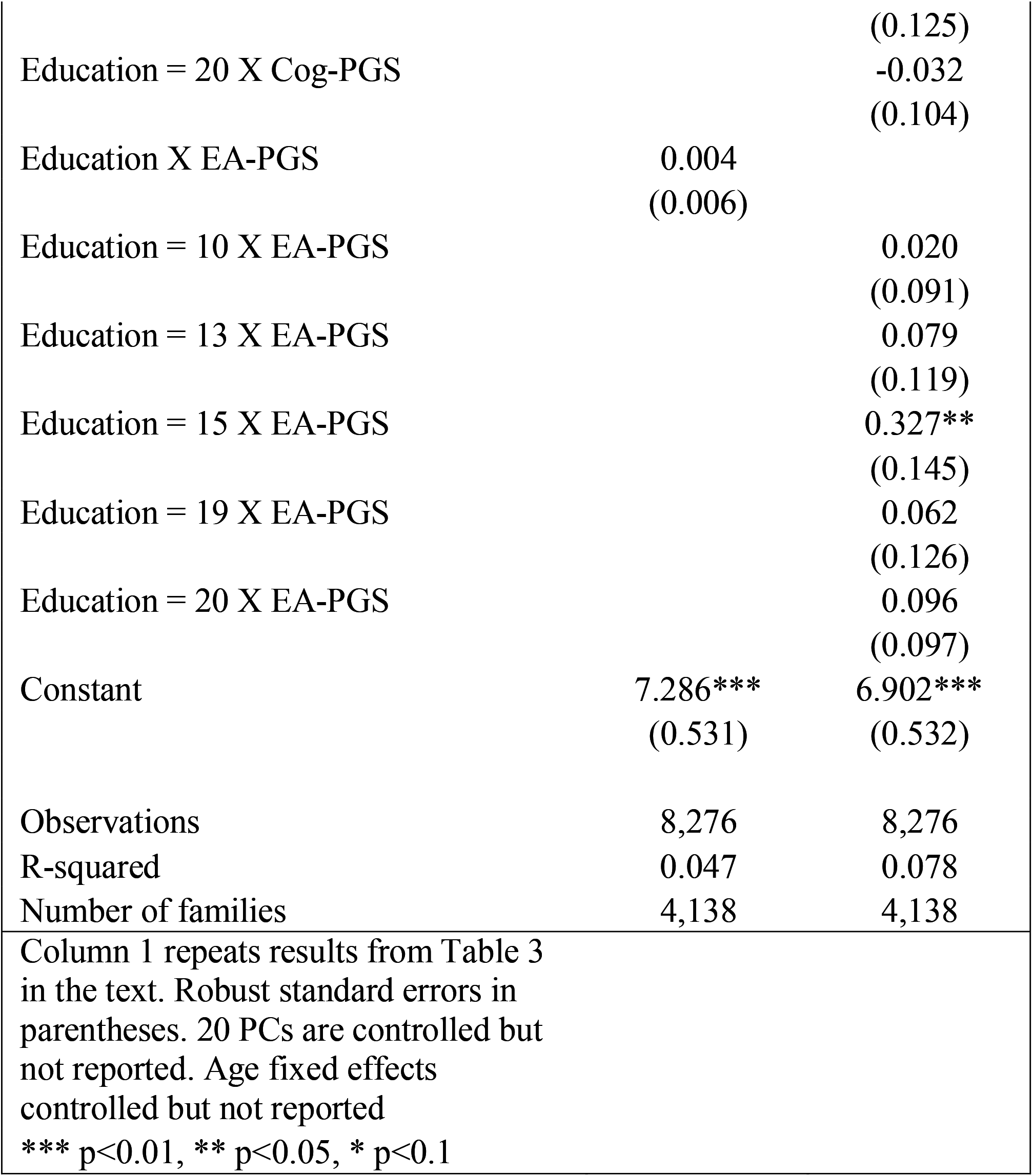
Exploring GxE across Educational Attainment Categories

## Footnotes

2 A study done by Fors and colleagues (2009) found that people who grew up with many siblings had less educational attainment later in life, but sibling number was not significantly associated with cognition in later life. The study did not look at differences between the siblings themselves.

3 These data are restricted, but one can gain access by following the procedures described in www.ukbiobank.ac.uk/register-apply/.

4 https://biobank.ndph.ox.ac.uk/showcase/field.cgi?id=20016

## References

Aartsen, M. J., Cheval, B., Sieber, S., Van der Linden, B. W., Gabriel, R., Courvoisier, D. S., Guessous, I., Burton-Jeangros, C., Blane, D., Ihle, A., Kliegel, M., & Cullati, S. (2019). Advantaged socioeconomic conditions in childhood are associated with higher cognitive functioning but stronger cognitive decline in older age. Proceedings of the National Academy of Sciences of the United States of America, 116(12), 5478–5486. https://doi.org/10.1073/pnas.1807679116

Ailshire, J., Karraker, A., & Clarke, P. (2017). Neighborhood social stressors, fine particulate matter air pollution, and cognitive function among older U.S. adults. Social Science & Medicine, 172, 56–63. https://doi.org/10.1016/j.socscimed.2016.11.019

Baier, T. (2019). Does sibling and twin similarity in cognitive ability differ by parents’ education? Journal of Family Research, 31(1).

Banks, J., & Mazzonna, F. (2012). The Effect of Education on Old Age Cognitive Abilities: Evidence from a Regression Discontinuity Design. Economic Journal, 122(560), 418– 448. https://doi.org/10.1111/j.1468-0297.2012.02499.x

Barnes, L. L., Leurgans, S., Aggarwal, N. T., Shah, R. C., Arvanitakis, Z., James, B. D., Buchman, A. S., Bennett, D. A., & Schneider, J. A. (2015). Mixed pathology is more likely in black than white decedents with Alzheimer dementia. Neurology, 85(6), 528– 534. https://doi.org/10.1212/WNL.0000000000001834

Boss, L., Kang, D.-H., & Branson, S. (2015). Loneliness and cognitive function in the older adult: a systematic review. International Psychogeriatrics / IPA, 27(4), 541–553. https://doi.org/10.1017/S1041610214002749

Chen, Y. Lv, C., Li, X., Zhang, J., Chen, K., Liu, Z., …Zhang, Z. (2019). The positive impacts of early-life education on cognition, leisure activity, and brain structure in healthy aging. Aging, 11(14), 4923–4942. https://doi.org/10.18632/aging.102088

Choi, S.W., & O’Reilly, P.F. (2019). PRSice-2: Polygenic Risk Score software for biobank-scale data. GigaScience, 8(7), giz082. https://doi.org/10.1093/gigascience/giz082

Clouston, S. A., Kuh, D., Herd, P., Elliott, J., Richards, M., & Hofer, S. M. (2012). Benefits of educational attainment on adult fluid cognition: international evidence from three birth cohorts. International Journal of Epidemiology, 41(6), 1729–1736. https://doi.org/10.1093/ije/dys148

Clouston, S. A., Smith, D. M., Mukherjee, S., Zhang, Y., Hou, M.W. Link, B.G., & Richards, M. (2020). Education and cognitive decline: an integrative analysis of global longitudinal studies of cognitive aging. The Journals of Gerontology: Series B, 75(7), e151–e160. https://doi.org/10.1093/geronb/gbz053

Cook, C. J., & Fletcher, J. M. (2015). Can education rescue genetic liability for cognitive decline? Social Science & Medicine, 127, 159–170. https://doi.org/10.1016/j.socscimed.2014.06.049

Davies, N. M., Dickson, M., Smith, G. D., van den Berg, G. B., & Windmeijer, F. (2018). The causal effects of education on health outcomes in the UK Biobank. Natural Human Behaviour, 2(2), 117–125. https://doi.org/10.1038/s41562-017-0279-y

Deary, I. J., & Johnson, W. (2010). Intelligence and education: causal perceptions drive analytic processes and therefore conclusions. International Journal of Epidemiology, 39(5), 1362– 1369. https://doi.org/10.1093/ije/dyq072

Dhuey, E., Figlio, D., Karbownik, K., & Roth, J. (2019). School Starting Age and Cognitive Development. Journal of Policy Analysis and Management: [the Journal of the Association for Public Policy Analysis and Management], 38(3), 538–578. https://doi.org/10.1002/pam.22135

Evans, I. E. M., Llewellyn, D. J., Matthews, F. E., Woods, R. T., Brayne, C., Clare, L., & CFAS-Wales research team. (2018). Social isolation, cognitive reserve, and cognition in healthy older people. PloS One, 13(8), e0201008. https://doi.org/10.1371/journal.pone.0201008

Fors, S., Lennartsson, C., & Lundberg, O. (2009). Childhood living conditions, socioeconomic position in adulthood, and cognition in later life: exploring the associations. The Journals of Gerontology. Series B, Psychological Sciences and Social Sciences, 64(6), 750–757. https://doi.org/10.1093/geronb/gbp029

Fletcher, J. M., Wu, Y., Zhao, Z., & Lu, Q. (2020). The production of within-family inequality: Insights and implications of integrating genetic data. In Genetics (No. biorxiv;2020.06.06.137778v1; p. 1883). bioRxiv. https://doi.org/10.1101/2020.06.06.137778

Galama, T., Lleras-Muney, A., & Kippersluis, H. V. (2018). The Effect of Education on Health and Mortality: A Review of Experimental and Quasi-Experimental Evidence. Oxford Research Encyclopedia of Economics and Finance. https://doi.org/10.1093/acrefore/9780190625979.013.7

Gill, T. M., Gahbauer, E. A., Han, L., & Allore, H. G. (2010). Trajectories of Disability in the Last Year of Life. The New England Journal of Medicine, 362(13), 1173–1180. https://doi.org/10.1056/NEJMoa0909087

Glymour, M. M., Kawachi, I., Jencks, C. S., & Berkman, L. F. (2008). Does childhood schooling affect old age memory or mental status? Using state schooling laws as natural experiments. Journal of Epidemiology and Community Health, 62(6), 532–537. https://doi.org/10.1136/jech.2006.059469

Glymour, M. M., Tzourio, C., & Dufouil, C. (2012). Is cognitive aging predicted by one’s own or one’s parents’ educational level? Results from the three-city study. American Journal of Epidemiology, 175(8), 750–759. https://doi.org/10.1093/aje/kwr509

Gow, A. J., Pattie, A., Whiteman, M. C., Whalley, L. J., & Deary, I. J. (2007). Social support and successful aging: Investigating the relationships between lifetime cognitive change and life satisfaction. Journal of Individual Differences, 28(3), 103–115. https://doi.org/10.1027/1614-0001.28.3.103

Howell, J. C., Watts, K. D., Parker, M. W., Wu, J., Kollhoff, A., Wingo, T. S., Dorbin, C. D., Qiu, D., & Hu, W. T. (2017). Race modifies the relationship between cognition and Alzheimer’s disease cerebrospinal fluid biomarkers. Alzheimer’s Research & Therapy, 9(1), 88. https://doi.org/10.1186/s13195-017-0315-1

Kuiper, J. S., Zuidersma, M., Zuidema, S. U., Burgerhof, J. G., Stolk, R. P., Oude Voshaar, R. C., & Smidt, N. (2016). Social relationships and cognitive decline: a systematic review and meta-analysis of longitudinal cohort studies. International Journal of Epidemiology, 45(4), 1169–1206. https://doi.org/10.1093/ije/dyw089

Kunkle, B.W., Grenier-Boley, B., Sims, R., Bis, J.C., Damotte, V., Naj, A.C., Boland, A., Vronskaya, M., Van Der Lee, S.J., Amlie-Wolf, A. and Bellenguez, C., 2019. Genetic meta-analysis of diagnosed Alzheimer’s disease identifies new risk loci and implicates Aβ, tau, immunity and lipid processing. Nature genetics, 51(3), pp.414–430. https://doi.org/10.1038/s41588-019-0358-2

Lee, J. J., Wedow, R., Okbay, A., Kong, E., Maghzian, O., Zacher, M., Nguyen-Viet, T. A., Bowers, P., Sidorenko, J., Karlsson Linnér, R., Fontana, M. A., Kundu, T., Lee, C., Li, H., Li, R., Royer, R., Timshel, P. N., Walters, R. K., Willoughby, E. A., … Social Science Genetic Association Consortium. (2018). Gene discovery and polygenic prediction from a genome-wide association study of educational attainment in 1.1 million individuals. Nature Genetics, 50(8), 1112–1121. https://doi.org/10.1038/s41588-018-0147-3

Lenehan, M. E., Summers, M. J., Saunders, N. L., Summers, J. J., & Vickers, J. C. (2015). Relationship between education and age-related cognitive decline: a review of recent research. Psychogeriatrics: The Official Journal of the Japanese Psychogeriatric Society, 15(2), 154–162. https://doi.org/10.1111/psyg.12083

Lövdén, M., Fratiglioni, L., Glymour, M. M., Lindenberger, U., & Tucker-Drob, E. M. (2020). Education and Cognitive Functioning Across the Life Span. Psychological Science in the Public Interest: A Journal of the American Psychological Society, 21(1), 6–41. https://doi.org/10.1177/1529100620920576

Luchetti, M., Terracciano, A., Stephan, Y., & Sutin, A. R. (2016). Personality and Cognitive Decline in Older Adults: Data from a Longitudinal Sample and Meta-Analysis. The Journals of Gerontology. Series B, Psychological Sciences and Social Sciences, 71(4), 591–601. https://doi.org/10.1093/geronb/gbu184

Meng, X., & D’Arcy, C. (2012). Education and dementia in the context of the cognitive reserve hypothesis: a systematic review with meta-analyses and qualitative analyses. PloS One, 7(6), e38268. https://doi.org/10.1371/journal.pone.0038268

Moorman, S. M., Carr, K., & Greenfield, E. A. (2018). Childhood socioeconomic status and genetic risk for poorer cognition in later life. Social Science & Medicine, 212, 219–226. https://doi.org/10.1016/j.socscimed.2018.07.025

Panza, F., Lozupone, M., Solfrizzi, V., Sardone, R., Dibello, V., Di Lena, L., D’Urso, F., Stallone, R., Petruzzi, M., Giannelli, G., Quaranta, N., Bellomo, A., Greco, A., Daniele, A., Seripa, D., & Logroscino, G. (2018). Different Cognitive Frailty Models and Health- and Cognitive-related Outcomes in Older Age: From Epidemiology to Prevention. Journal of Alzheimer’s Disease, 62(3), 993–1012. https://doi.org/10.3233/JAD-170963

Polderman, T. J. C., Benyamin, B., de Leeuw, C. A., Sullivan, P. F., van Bochoven, A., Visscher, P. M., & Posthuma, D. (2015). Meta-analysis of the heritability of human traits based on fifty years of twin studies. Nature Genetics, 47(7), 702–709. https://doi.org/10.1038/ng.3285

Plassman, B. L., Williams, J. W., Jr, Burke, J.R., Holsinger, T., & Benjamin, S. (2010). Systematic review: factors associated with risk for and possible prevention of cognitive decline in later life. Annals of Internal Medicine, 153(3), 182–193. https://doi.org/10.7326/0003-4819-153-3-201008030-00258

Reynolds, C. A., Finkel, D., & Zavala, C. (2014). Gene by Environment Interplay in Cognitive Aging. In D. Finkel & C. A. Reynolds (Eds.), Behavior Genetics of Cognition Across the Lifespan (pp. 169–199). Springer New York. https://doi.org/10.1007/978-1-4614-7447-0_6

Richards, M., & Hatch, S. L. (2011). A life course approach to the development of mental skills. The Journals of Gerontology. Series B, Psychological Sciences and Social Sciences, 66 Suppl 1, i26–i35. https://doi.org/10.1093/geronb/gbr013

Richards, M. & Sacker, A. (2011). Is education causal? Yes. International Journal of Epidemiology, 40(2), 516–518. https://doi.org/10.1093/ije/dyq166

Rietveld, C.A., Esko, T., Davies, G., Pers, T.H., Turley, P., Benyamin, B., Chabris, C.F., Emilsson, V., Johnson, A.D., Lee, J.J. and De Leeuw, C., 2014. Common genetic variants associated with cognitive performance identified using the proxy-phenotype method. Proceedings of the National Academy of Sciences, 111(38), pp.13790–13794. https://doi.org/10.1073/pnas.1404623111

Ritchie, S. J., Tucker-Drob, E. M., Cox, S. R., Corley, J., Dykiert, D., Redmond, P., Pattie, A., Taylor, A. M., Sibbett, R., Starr, J. M., & Deary, I. J. (2016). Predictors of ageing-related decline across multiple cognitive functions. Intelligence, 59, 115–126. https://doi.org/10.1016/j.intell.2016.08.007

Robertson, D. A., Savva, G. M., & Kenny, R. A. (2013). Frailty and cognitive impairment--a review of the evidence and causal mechanisms. Ageing Research Reviews, 12(4), 840– 851. https://doi.org/10.1016/j.arr.2013.06.004

Schneeweis, N., Skirbekk, V., & Winter-Ebmer, R. (2014). Does education improve cognitive performance four decades after school completion? Demography, 51(2), 619–643. https://doi.org/10.1007/s13524-014-0281-1

Sofi, F., Valecchi, D., Bacci, D., Abbate, R., Gensini, G. F., Casini, A., & Macchi, C. (2011). Physical activity and risk of cognitive decline: a meta-analysis of prospective studies. Journal of Internal Medicine, 269(1), 107–117. https://doi.org/10.1111/j.1365-2796.2010.02281.x

Stern, Y. (2009). Cognitive reserve. Neuropsychologia, 47(10), 2015–2028. https://doi.org/10.1016/j.neuropsychologia.2009.03.004

Stern, Y. (2012). Cognitive reserve in ageing and Alzheimer’s disease. The Lancet Neurology, 11(11), 1006–1012. https://doi.org/10.1016/S1474-4422(12)70191-6

van den Kommer, T. N., Comijs, H. C., Aartsen, M. J., Huisman, M., Deeg, D. J. H., & Beekman, A. T. F. (2013). Depression and Cognition: How Do They Interrelate in Old Age? The American Journal of Geriatric Psychiatry: Official Journal of the American Association for Geriatric Psychiatry, 21(4), 398–410. https://doi.org/10.1016/j.jagp.2012.12.015

Vossius, C., Larsen, J. P., Janvin, C., & Aarsland, D. (2011). The economic impact of cognitive impairment in Parkinson’s disease. Movement Disorders: Official Journal of the Movement Disorder Society, 26(8), 1541–1544. https://doi.org/10.1002/mds.23661

Wilson, R. S., Yu, L., Lamar, M., Schneider, J. A., Boyle, P. A., & Bennett, D. A. (2019). Education and cognitive reserve in old age. Neurology, 92(10), e1041–e1050. https://doi.org/10.1212/WNL.0000000000007036

Yount, K. M. (2008). Gender, resources across the life course, and cognitive functioning in Egypt. Demography, 45(4), 907–926. https://doi.org/10.1353/dem.0.0034

Zahodne, L. B., Glymour, M. M., Sparks, C., Bontempo, D., Dixon, R. A., MacDonald, S. W. S., & Manly, J. J. (2011). Education does not slow cognitive decline with aging: 12-year evidence from the Victoria longitudinal study. Journal of the International Neuropsychological Society, 17(6), 1039–1046. https://doi.org/10.1017/S1355617711001044

Zaninotto, P., Batty, G. D., Allerhand, M., & Deary, I. J. (2018). Cognitive function trajectories and their determinants in older people: 8 years of follow-up in the English Longitudinal Study of Ageing. Journal of Epidemiology and Community Health, 72(8), 685–694. https://doi.org/10.1136/jech-2017-210116

